# Association between ABO Blood Group and COVID-19 Pneumonia Severity: A Cross-Sectional Study from Ethiopia’s First Designated COVID-19 Treatment Center

**DOI:** 10.64898/2026.09.04.26362275

**Authors:** Eden Bekele Urgecha, Fanuel Alemayehu Tefera, Zewdie Aderaw Alemu, Solomon Teshome Mekonen, Etsub Bekele Orgecha, Saron Negasi Gidey, Fiyameta Abraham Fissehatsion

## Abstract

**Background:** Coronavirus disease 2019 (COVID-19), caused by severe acute respiratory syndrome coronavirus 2, spreads rapidly and can cause severe acute respiratory failure; advanced age, chronic disease, and other comorbidities increase the risk of severe infection. By early 2022, more than 477 million cases and 6.11 million deaths had been reported globally. This study assessed the association between ABO blood group and severity of COVID-19 pneumonia among patients admitted to Eka Kotebe General Hospital, Addis Ababa, Ethiopia.

**Methods:** A hospital-based cross-sectional study was conducted using systematic random sampling; data were abstracted from patient records, and severity was modeled using ordinal logistic regression, with multicollinearity and the proportional odds assumption checked via the variance inflation factor and test of parallel lines, and model fit assessed by likelihood ratio chi-square and goodness-of-fit tests.

**Results:** Of 355 patients included (100% response rate), 54.9% were male and 31.0% were older than 61 years (median age, 48 years; interquartile range, 33-65). Overall, 10.14% were asymptomatic, 24.79% had mild/moderate disease, 60.56% had severe disease, and 4.51% had critical disease. Pregnancy and presence of complications were significantly associated with severity (adjusted odds ratio, 0.15 [95% CI, 0.04-0.57] and 14.96 [95% CI, 4.14-54.01], respectively); ABO blood group showed no association with severity.

**Conclusion:** These findings demonstrate a substantial association between COVID-19 severity and complications and pregnancy, but not ABO blood group.

## 1. Introduction

Coronavirus disease 2019 (COVID-19) is defined as illness caused by a novel coronavirus called severe acute respiratory syndrome coronavirus 2 (SARS-CoV-2), which was first identified amid an outbreak of respiratory illness cases in Wuhan City, Hubei Province, China, and was initially reported to the WHO on December 31, 2019. On March 11, 2020, the WHO declared COVID-19 a global pandemic, its first such designation since declaring H1N1 influenza a pandemic in 2009. Exposure to respiratory droplets carrying infectious virus and airborne transmission of droplets are the main stay for transmission of infection. [1]

Presentations of COVID-19 range from asymptomatic/mild symptoms to severe illness and mortality. Complications include pneumonia, acute respiratory distress syndrome, cardiac injury, arrhythmia, septic shock, liver dysfunction, acute kidney injury, and multi-organ failure, among others. Approximately 5% of patients with COVID-19, and 20% of those hospitalized, experience severe symptoms necessitating intensive care, and ICU case fatality is reported up to 40%. [1,2]

The CDC has released a summary of evidence-based comorbidities, supported by meta-analysis and systematic review, that carry a strong correlation with risk of severe COVID-19 illness. These include cancer, cerebrovascular disease, chronic kidney disease, COPD, diabetes mellitus, heart conditions, immune-compromised state from solid organ transplant, obesity, pregnancy, and smoking. Apart from comorbidities, old age and male sex tend to be associated with more severe COVID-19 illness. [2]

On the surface of red blood cells, human ABO blood type antigens display diverse phenotypes and genetically produced glycoconjugate structures that are actively involved in the physiology and pathophysiology of the cells. Since it was discovered that antibodies and antigens are inherited, associations between blood type and disease have been researched extensively. Early etiological studies indicated that blood type O has a connection with increased incidence of cholera, plague, tuberculosis, and mumps, whereas blood type A is linked with increased incidence of smallpox and Pseudomonas aeruginosa infection; blood type B is associated with increased incidence of gonorrhea, tuberculosis, Streptococcus pneumoniae, E. coli, and salmonella infections; and blood type AB is associated with increased incidence of smallpox, E. coli, and salmonella infections. The link between ABO blood type and thromboembolic disease and bleeding risk is mediated by glycosyltransferase activity and the plasma level and biologic activity of von Willebrand factor, a carrier protein for coagulation factor VIII that is comparatively low in blood type O. [3]

Reported prevalence in Ethiopia has been considerably lower than global figures. A community-based cross-sectional study conducted in Ataye, Northeast Ethiopia, involving 8,752 participants found an overall prevalence of 3.3%. A retrospective cross-sectional study in West Gondar zone, Northwest Ethiopia, enrolling 1,166 participants, found a prevalence of 1.37% (95% CI: 0.66–2.08), with living in an urban area and being female statistically associated with a positive result. A separate seroprevalence study estimated IgG prevalence in Addis Ababa at 1.9% (95% CI 0.4–3.7%), with combined IgM/IgG prevalence at 3.5% (95% CI 1.7–5.4%), and higher seroprevalence in central than peripheral sub-cities. [4,5]

### ABO blood group and COVID-19 susceptibility

A multi-institutional study in the USA found that, of patients testing positive, 34.2% were blood type A, 15.6% type B, 4.7% type AB, and 45.5% type O; blood type A had no correlation with positive testing, whereas types B and AB were associated with higher odds of testing positive and type O with a lower risk. [1] A retrospective analysis in Turkey similarly found blood group A significantly over-represented (57%) among COVID-19 patients, with blood group O significantly under-represented (24.8%), although blood group type did not affect clinical outcome once patients were infected. [6] A retrospective study in Kuwait found that, of positive patients, 37.1%, 25.5%, 28.9%, and 8.5% were groups O, A, B, and AB respectively, a lower frequency of group O and a higher frequency of B and AB compared with the general population. [7] By contrast, a study from Nigeria found the blood group distribution, 56.9% group O, 23.5% group A, 19.6% group B, and no group AB positive patients, to be comparable to the general population, with no relationship found between ABO blood group and COVID-19 positivity. [8] A study from Tunisia found a statistically significant higher prevalence of blood group A among COVID-19 patients compared with blood donors and disease-free controls, [9] while a case-control study from Sudan found that Rh-positive, but not Rh-negative, individuals were more vulnerable to infection, and that O-positive individuals were least exposed to severe symptoms. [10]

### ABO blood group and COVID-19 severity

A population-based cohort study in Canada reported that blood group O may be associated with a slightly lower risk of COVID-19 and severity of illness, with type B at significantly higher risk for severe illness than type A. [11] A literature review similarly suggested blood type O may serve as a protective factor, with individuals of non-A blood types, specifically O or B, found to be less susceptible to infection, [12] a pattern echoed by another study suggesting group O carries a lower risk of infection and group A a higher risk of infection along with severe disease. [13] A cohort study in Austria suggested blood types A, B, and O to possess protective factors, whereas group AB was a risk factor for COVID-19, [14] and a literature review from Brazil proposed that the protective effect of group O could be mediated either by natural anti-A and anti-B antibodies or by lower efficiency of furin cleavage in group O individuals. [15] A systematic review from Italy found low to very low evidence that group O individuals are less susceptible to infection compared with non-O individuals, with no evidence of an effect of blood type on disease severity. [16]

In contrast, a cohort study in China concluded that, although the prevalence of blood group O was higher in the cohort, hospital stay, severity of disease, and mortality were all associated with blood group A, [17] a finding corroborated by a systematic review and meta-analysis from the same country reporting that blood group A carried a substantially higher risk of both COVID-19 infection and mortality, with blood group O not prone to develop the disease. [18]

### ABO blood group and COVID-19 outcome

A retrospective study in Iran involving 1,554 confirmed cases found that patients with type B− had 3.8 times higher odds of death than the A+ reference group, while types B+, AB+, O+, and O− all had lower odds of death than A+. [19] An observational study from the New York Presbyterian hospital system reported that blood type A carried a decreased risk of both intubation and death relative to type O, while type AB carried an increased risk of both outcomes, and type B carried a higher risk of intubation but a lower risk of death compared with type O. [13] A retrospective study in South Asia found an overall COVID-19-related mortality of 10.6%, highest in blood group A (13.9%), with no statistically significant difference in severity between groups. [6] A retrospective cohort study in the ICU of a tertiary COVID-dedicated hospital in Bangladesh found that patients with blood type A had a greater need for supplemental oxygen and were significantly more likely to die than patients with other blood types, after adjusting for age, sex, and non-communicable disease, indicating that blood type A carries a higher chance of death and other complications. [17] A single-center longitudinal study of Arab patients at Al-Amal Hospital found blood groups AB and B significantly associated with need for respiratory support and in-hospital death, respectively, compared with blood group O. [20] A study from Nigeria, however, found no statistically significant association between ABO blood group and COVID-19 severity or mortality, and a single-center study of Black patients similarly found no association between blood type and in-hospital mortality or ICU admission, although blood type A patients had a higher propensity for kidney injury that did not translate into worse survival. [8] A separate genetic association study similarly found no significant relationship between ABO blood group and either COVID-19 test positivity or mortality in men or women, nor any association with the ABO gene variant rs657152. [23]

### Other factors associated with COVID-19 severity

Socio-demographic factors have been repeatedly linked to COVID-19 severity: a cross-sectional analysis in Iran found the highest rates of low blood-oxygen saturation and death among patients aged 80 and older and among males; [21] a case-control study in the United States similarly found hospitalization and ICU admission associated with male sex, and viral positivity associated with non-White race.

Comorbidities are similarly implicated: a retrospective cohort of 167,500 individuals in Ontario, Canada found mortality risk rising from 2.14 to 4.81 times as the number of comorbidities increased from one to five or more, with the strongest predictors being solid organ transplant, dementia, chronic kidney disease, severe mental illness, cardiovascular disease, diabetes, COPD, cancer, and hypertension; [24] a literature search in Iran found comorbidities in 41% of patients, most strongly cerebrovascular disease; [25] and a cross-sectional study in eastern Ethiopia found more than half (52.4%) of patients had comorbid conditions, with diabetes, hypertension, cardiovascular disease, and marital status significant predictors of severe outcome. [26]

### Rationale of the study

There remains a paucity of published data on the effect of ABO blood group on the severity of COVID-19 pneumonia in Ethiopia. Even though more than two years have passed since the COVID-19 outbreak and the report of its entry into Ethiopia, only a handful of studies have examined the severity of COVID-19 pneumonia in the country, and no published study to date has specifically examined the association between blood group and COVID-19 severity in this setting. Establishing this relationship can aid in prioritizing individuals for vaccination at times when supply is scarce, enable clinicians to forecast likely prognosis and initiate aggressive treatment earlier, and inform prevention strategies to reduce the burden of the disease in the community and the country at large.

This study therefore aimed to assess the association between ABO blood group and severity of COVID-19 pneumonia among patients admitted to Eka Kotebe General Hospital. The specific objectives were to determine the prevalence of COVID-19 severity among admitted patients, to identify factors associated with severity of COVID-19 pneumonia, and to investigate differences in outcome among COVID-19 patients with different blood groups.

## 2. Materials and Methods

### 2.1. Study area and period

Eka Kotebe General Hospital, COVID-19 Isolation and Treatment Center, located in Addis Ababa, Ethiopia, was the first hospital designated to manage confirmed COVID-19 cases in the country, with a capacity to admit 600 cases. The study was conducted from January 2021 to January 2022.

### 2.2. Study design

A hospital-based cross-sectional study was conducted at Eka Kotebe General Hospital, COVID-19 Isolation and Treatment Center.

### 2.3. Source and study population

The source population was all COVID-19 patients admitted to Eka Kotebe General Hospital, COVID-19 Isolation and Treatment Center. The study population was all COVID-19 patients admitted with a diagnosis of COVID-19 pneumonia who fulfilled the inclusion criteria during the study period (January 2021 - January 2022).

### 2.4. Sample size and sampling procedure

The sample size was determined using the single population proportion formula, taking the largest proportion from the literature review (0.36, used as 0.3 in the calculation), a 95% confidence level, and a 5% margin of error, yielding an initial size of 323; after adding a 10% non-response allowance, the final sample size was 355. A sample size was also computed using the double population proportion formula for the exposed/unexposed comparison, yielding a substantially smaller size of 56 at 80% power; because this was markedly smaller, the sample size from the single population proportion formula (355) was used.

Systematic random sampling was employed to select eligible participants over the course of one month. With approximately 10,000 patients admitted to the hospital during the sampling frame and a target sample size of 355, the sampling interval was 28; every 28th patient’s medical record, ordered by arrival time to the emergency room and fulfilling the inclusion and exclusion criteria, was assessed.

### 2.5. Inclusion and exclusion criteria

Inclusion criteria: all patients admitted to the hospital who tested positive for COVID-19 by RDT within the study period.

Exclusion criteria: patients discharged against medical advice, having signed full responsibility for denial of treatment.

### 2.6. Study variables

The dependent variable was COVID-19 clinical severity (mild/moderate, severe, critical). Independent variables included ABO blood group; socio-demographic factors (sex, age, marital status, occupation, residency); comorbidities (pregnancy, diabetes mellitus, hypertension, malignancy, cerebrovascular accident, chronic kidney disease, cardiac illness, COPD, HIV, TB, chronic liver disease, and others); presence of complications; events during hospital stay; and duration of illness.

### 2.7. Operational definitions

Asymptomatic/pre-symptomatic infection: individuals testing positive for SARS-CoV-2 by a virologic test but without symptoms consistent with COVID-19. Mild illness: presence of any COVID-19 signs or symptoms without dyspnea, with normal oxygen saturation and normal chest imaging. Moderate illness: mild disease with evidence of lower respiratory involvement on assessment or imaging and SpO2 ≥94% on room air. Severe illness: moderate disease with SpO2 <94% on room air at sea level, PaO2/FiO2 ratio <300 mmHg, respiratory rate >30/min, or lung infiltrates >50%. Critical illness: severe disease with intubation for respiratory failure, septic shock, and/or multi-organ dysfunction.

### 2.8. Data collection and quality assurance

A structured, English-language data collection checklist was developed based on review of records and similar literature, and pretested at Eka Kotebe General Hospital prior to the study period, with amendments made as needed. Five general practitioners and a nurse received an hour of training on the checklist and data entry procedures. Records of patients admitted between January 2021 and January 2022 were reviewed, with data collected in April 2022 and checked for consistency and completeness before entry and analysis.

### 2.9. Data management and analysis

Data were coded and entered into Epi Info, cleaned, and exported to SPSS version 27.0 for analysis. Categorical variables were summarized using frequencies and percentages, and numerical variables using medians and interquartile ranges. The association between independent variables and severity of COVID-19 was assessed using an ordinal logistic regression model. Univariate ordinal logistic regression was run at a 25% level of significance to screen candidate variables, and multivariate ordinal logistic regression was then run including the significant univariate variables. Adjusted odds ratios (AOR), p-values, and 95% confidence intervals were used to assess the strength of association, with variables at p ≤ 0.05 in the final model considered significantly associated with the outcome.

Model assumptions were checked using the variance inflation factor (VIF < 10, indicating no meaningful multicollinearity) and the test of parallel lines (p = 0.279, satisfying the proportional odds assumption). The final multivariate model showed adequate fit, with p = 0.000 for the likelihood ratio chi-square test and p = 0.109 and p = 1.000 for the Pearson and deviance goodness-of-fit tests, respectively.

### 2.10. Ethical consideration

Ethical clearance for this study was obtained from the Ethical Review Committee of Eka Kotebe General Hospital, Addis Ababa, Ethiopia (approval date: 08 January 2021; reference number: EKGH/IRERC/027/2021). The committee reviewed the study protocol and found no violation of basic research ethics principles in either the methodology of data acquisition or the overall content, and authorized the study to proceed at the hospital. Patient information was collected using a structured tool, kept entirely anonymous, and used only for the intended purpose of the study, with confidentiality assured throughout. Because the study relied on retrospective review of de-identified medical records, the committee waived the requirement for individual informed consent.

## 3. Results

### 3.1. Socio-demographic characteristics

A total of 355 study subjects were involved, with a response rate of 100%. More than half (54.9%) of respondents were male. Nearly one-third (31.0%) of participants were older than 61 years, with an overall median age of 48 years (IQR: 33–65 years); ages ranged from 18 to 92 years. The majority (90.4%) of subjects were urban residents (Table 1).

**Table 1:**
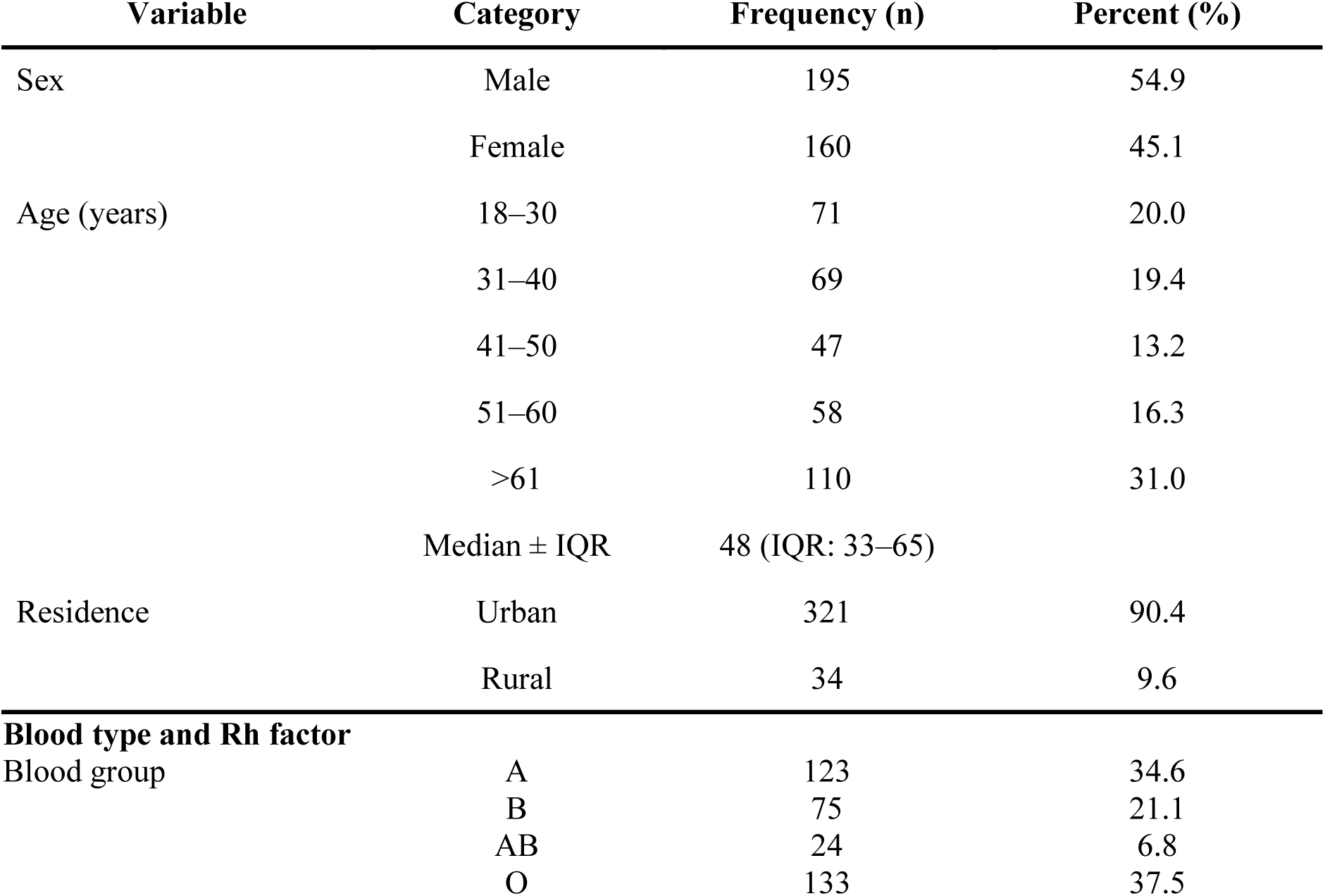
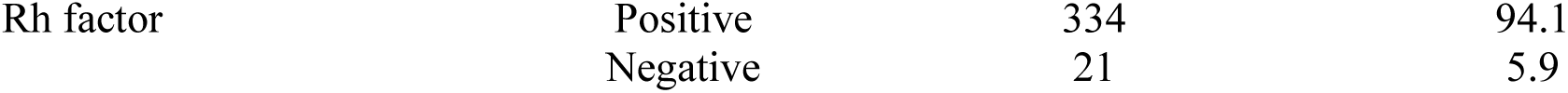
Socio-demographic characteristics and blood type of study participants admitted at Eka Kotebe General Hospital, COVID-19 Isolation and Treatment Center, 2022.

### 3.2. Blood type and Rh factor

More than one-third (37.5%) of subjects had blood group O and 34.6% had blood group A. The majority (94.1%) were Rh-positive (Table 1).

### 3.3. Symptoms at admission

89.3% of participants had symptoms at admission; of these, 73.5% had fever, 92.7% had cough, 12.6% had sore throat, 18.3% had nausea/vomiting, 9.5% had diarrhea, 13.9% had loss of taste/smell, 44.5% had shortness of breath, and 11.4% had other symptoms (Table 2).

**Table 2:** Symptoms and comorbidities at admission among study participants admitted at Eka Kotebe General Hospital, COVID-19 Isolation and Treatment Center, 2022.

| Symptom | Response | Frequency (n) | Percent (%) |
| --- | --- | --- | --- |
| Any symptom | Yes | 317 | 89.3 |
| Fever | Yes | 233 | 73.5 |
| Cough | Yes | 294 | 92.7 |
| Sore throat | Yes | 40 | 12.6 |
| Nausea/vomiting | Yes | 58 | 18.3 |
| Diarrhea | Yes | 30 | 9.5 |
| Loss of taste/smell | Yes | 44 | 13.9 |
| Shortness of breath | Yes | 141 | 44.5 |
| Other | Yes | 36 | 11.4 |
| **Comorbidities** |  |  |  |
| Any comorbidity | Yes | 220 | 62.0 |
| Pregnancy | Yes | 52 | 23.6 |
| Diabetes mellitus | Yes | 90 | 40.9 |
| Hypertension | Yes | 100 | 45.5 |
| Malignancy | Yes | 7 | 3.2 |
| CVA / stroke | Yes | 8 | 3.6 |
| Renal disease (CKD) | Yes | 12 | 5.5 |
| Cardiac illness (HF, CAD, CMP) | Yes | 18 | 8.2 |
| COPD (incl. asthma) | Yes | 22 | 10.0 |
| HIV | Yes | 16 | 7.3 |
| TB | Yes | 3 | 1.4 |
| Chronic liver disease | Yes | 4 | 1.8 |
| Other (smoking, substance use, obesity, etc.) | Yes | 22 | 10.0 |

### 3.4. Comorbidities

62.0% of participants had at least one comorbidity. Hypertension (45.5%) and diabetes mellitus (40.9%) were the most common, followed by COPD/asthma (10.0%), other conditions including smoking/substance use (10.0%), cardiac illness (8.2%), HIV (7.3%), renal disease (5.5%), CVA/stroke (3.6%), obesity (3.1%), malignancy (3.2%), chronic liver disease (1.8%), and TB (1.4%). Pregnancy was present in 23.6% of participants (Table 2).

### 3.5. Severity and outcome of COVID-19 infection

10.14% of participants were asymptomatic, 24.79% were in the mild/moderate stage, 60.56% were in the severe stage, and 4.51% were in the critical stage of COVID-19 infection (Figure 1).

**Figure 1:**
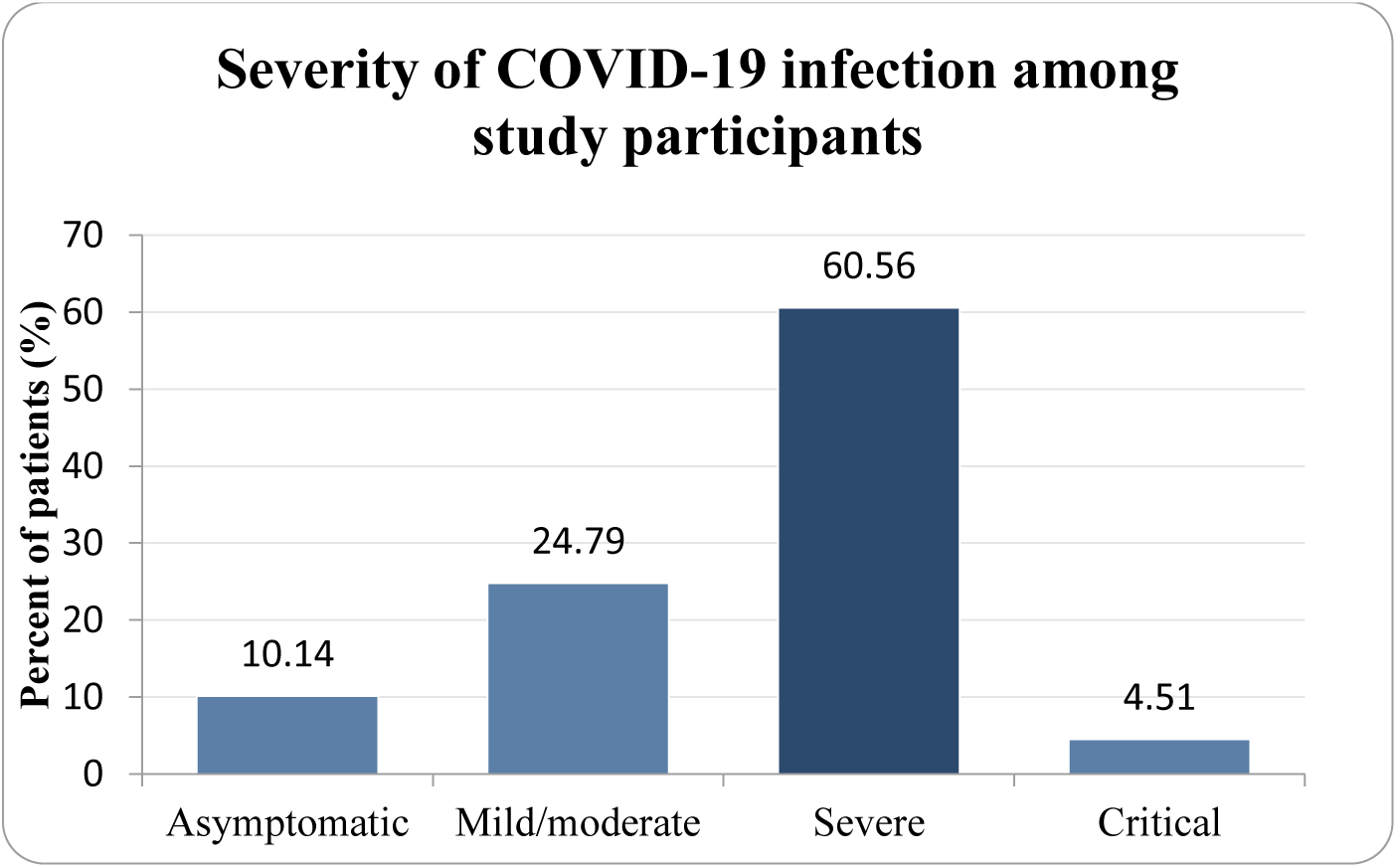
Severity of COVID-19 infection among study participants admitted at Eka Kotebe General Hospital, COVID-19 Isolation and Treatment Center, 2022. *Alt text: Bar chart of COVID-19 severity among 355 participants, showing 10.14% asymptomatic, 24.79% mild/moderate, 60.56% severe, and 4.51% critical*.

Regarding oxygen requirement and complications (Table 3), 35.2% of participants required room air and 42.8% required intranasal oxygen at arrival to the emergency room. Of the 23.7% of subjects who developed complications, 27.4% had ARDS, 57.1% had sepsis/septic shock, and 75.0% had acute kidney injury. Overall, 27.0% required ICU admission and 18.0% required intubation; mean hospital stay was 13.89 ± 9.89 days. 66.20% of participants recovered, 22.25% died, and 11.55% were transferred to other health facilities (Figure 2).

**Figure 2:**
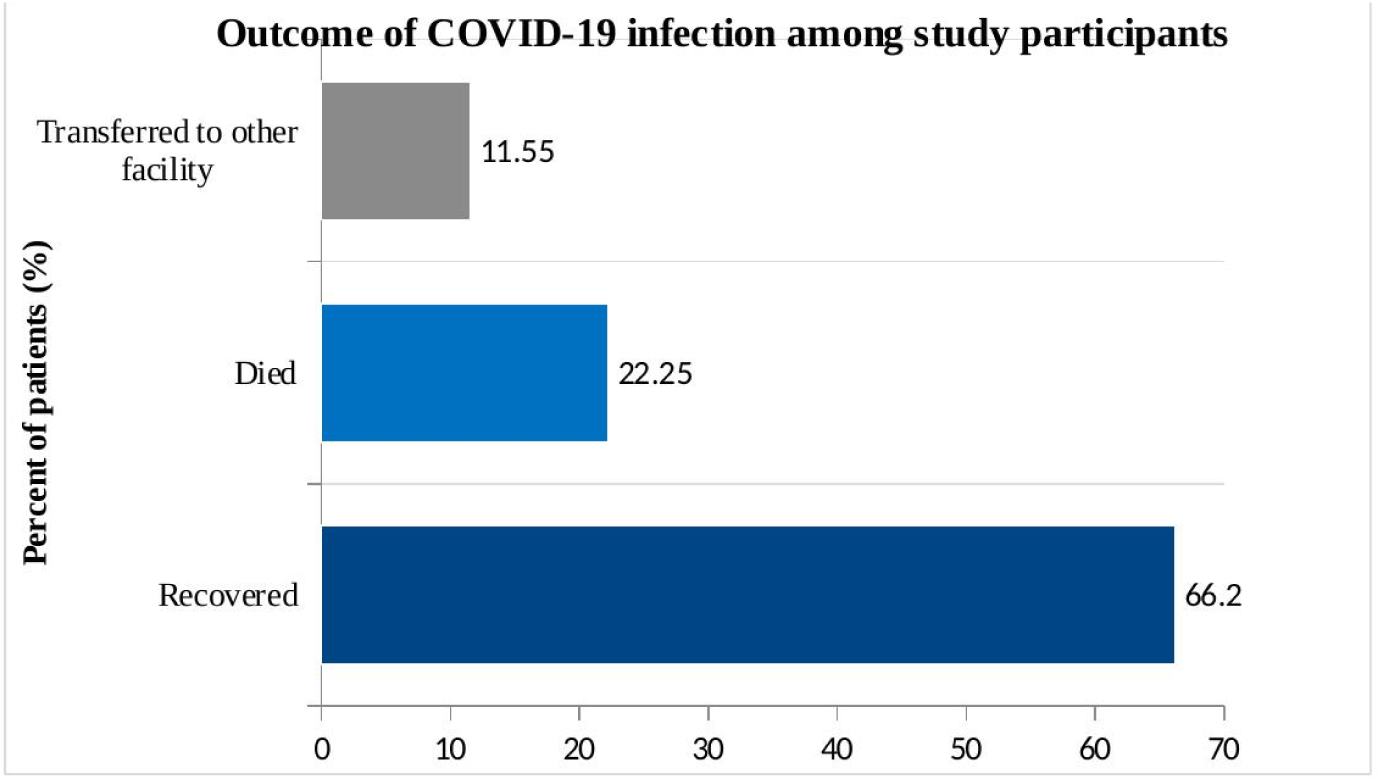
Outcome of COVID-19 infection among study participants admitted at Eka Kotebe General Hospital, COVID-19 Isolation and Treatment Center, 2022. *Alt text: Horizontal bar chart of COVID-19 outcomes among 355 participants, showing 66.2% recovered, 22.25% died, and 11.55% transferred to another facility*.

**Table 3:** Oxygen requirement at arrival and complications among study participants at Eka Kotebe General Hospital, COVID-19 Isolation and Treatment Center, 2022.

| Item | Response | Frequency | Percent |
| --- | --- | --- | --- |
| Oxygen at arrival | Room air | 125 | 35.2 |
|  | Intranasal oxygen | 152 | 42.8 |
|  | Simple face mask | 43 | 12.1 |
|  | Face mask w/ reservoir | 27 | 7.6 |
|  | CPAP | 7 | 2.0 |
|  | On mechanical ventilation | 1 | 0.3 |
| Any complication | Yes | 84 | 23.7 |
| ARDS | Yes (of complicated) | 23 | 27.4 |
| Sepsis / septic shock | Yes (of complicated) | 48 | 57.1 |
| AKI | Yes (of complicated) | 63 | 75.0 |
| Cardiac injury (MI) | Yes (of complicated) | 3 | 3.6 |
| Acute liver injury | Yes (of complicated) | 5 | 6.0 |
| Neurologic manifestation | Yes (of complicated) | 5 | 6.0 |
| Multi-organ failure | Yes (of complicated) | 6 | 7.1 |
| ICU admission | Yes | 96 | 27.0 |
| Intubation | Yes | 64 | 18.0 |
| Hospital stay (days) | Mean ± SD | 13.89 ± 9.89 | |

### 3.6. Univariable analysis

In univariable ordinal logistic regression (Table 4), sex, age group, symptom status, comorbidity, complication, pregnancy, ALT, BUN, creatinine, and length of hospital stay were statistically significant at the 25% level and were retained for the multivariable model. Residency, blood group, Rh factor, WBC, hemoglobin, platelet, ALP, and AST were not significant at this level and were excluded.

**Table 4:** Univariable and multivariable ordinal logistic regression analysis of factors associated with COVID-19 severity, using three ordered categories (Nagelkerke’s R^2^ = 0.432 for the multivariable model)

| Variable | Category | COR (95% CI) | P-value | AOR (95% CI) | P-value |
| --- | --- | --- | --- | --- | --- |
| Sex | Male | 1.98 (1.29, 3.04) | 0.002 | 0.54 (0.24, 1.22) | 0.140 |
|  | Female | 1 (ref) |  | - | - |
| Age (years) | 18–30 | 0.06 (0.03, 0.12) | <0.001 | 0.46 (0.12, 1.83) | 0.270 |
|  | 31–40 | 0.16 (0.08, 0.33) | <0.001 | 0.42 (0.14, 1.28) | 0.125 |
|  | 41–50 | 0.65 (0.29, 1.44) | 0.287 | 0.61 (0.22, 1.69) | 0.344 |
|  | 51–60 | 0.39 (0.19, 0.80) | 0.011 | 0.67 (0.23, 1.90) | 0.449 |
|  | >61 | 1 (ref) |  | - | - |
| Symptom | Yes | 29.97 (8.98, 100) | <0.001 | 2.79 (0.50, 15.67) | 0.245 |
| Comorbidity | Yes | 1.83 (1.18, 2.82) | 0.006 | - | - |
| Complication | Yes | 19.27 (7.97, 46.61) | <0.001 | 14.96 (4.14, 54.01) | <0.001 |
| Pregnancy | Yes | 0.09 (0.05, 0.19) | <0.001 | 0.15 (0.04, 0.57) | 0.005 |
| ALT | Mean ± SD | 1.002 (0.9996, 1.004) | 0.104 | 0.999 (0.996, 1.003) | 0.724 |
| BUN | Mean ± SD | 1.03 (1.01, 1.04) | <0.001 | 1.002 (0.98, 1.02) | 0.839 |
| Creatinine | Mean ± SD | 1.07 (0.98, 1.16) | 0.116 | 0.96 (0.87, 1.07) | 0.457 |
| Length of stay | Mean ± SD | 1.03 (1.004, 1.05) | 0.023 | 1.02 (0.99, 1.06) | 0.226 |

### 3.7. Multivariable analysis

Ten of eighteen candidate variables were retained for the multivariable ordinal logistic regression model based on their crude association at the 25% level of significance. No meaningful multicollinearity was present (VIF < 10), and the proportional odds assumption was satisfied (parallel-line test, p = 0.279). The final model showed adequate fit (likelihood ratio chi-square p = 0.000; Pearson goodness-of-fit p = 0.109; deviance goodness-of-fit p = 1.000).

The presence of complications was significantly associated with severity: patients with complications were 14.96 times more likely to be in a more severe category than those without, holding other variables constant (AOR = 14.96, 95% CI: 4.14–54.01, p < 0.001). Pregnancy was also significantly associated with severity (AOR = 0.15, 95% CI: 0.04–0.57, p = 0.005), indicating that pregnant patients were 85% less likely to be in a more severe category compared with non-pregnant patients (Table 4). No other variable, including ABO blood group, retained statistical significance in the adjusted model.

## 4. Discussion

At the time this study was conducted, COVID-19 remained a significant cause of hospitalization and death worldwide. This study used an ordinal logistic regression model to assess the association between ABO blood group and severity of COVID-19 pneumonia, and to identify other factors associated with severity, among 355 patients admitted to Eka Kotebe General Hospital.

First, ABO blood group was not associated with severity of clinical status in this cohort. The ABO blood type, found on chromosome 9 of human DNA and among the most frequently used typing systems in clinical practice, has been implicated in numerous infectious and non-infectious disorders in prior research. However, unlike the study conducted at New York Presbyterian, [13] no association between ABO blood group and severity was found in this single-institution, retrospective investigation. On this basis, ABO blood typing should not currently be regarded as a prognostic indicator in patients with COVID-19 in this setting.

The severity distribution observed in this cohort was broadly consistent with a study from Egypt reporting 38.3% of subjects with severe or critical disease, [27] though lower than a US study reporting 65% with moderate/severe and 35% with critical disease, a difference that may reflect differing socio-demographic characteristics, study design, and setting.

The recovery, mortality, and transfer rates observed reflect the substantial burden of the disease in this setting.

Among the 355 participants, 63.4% with blood type A, 58.7% with type B, 50% with type AB, and 60.9% with type O developed severe COVID-19 infection. This finding is consistent with a study from Bangladesh that similarly found blood type A significantly associated with greater disease severity and mortality, [17] and with a systematic review and meta-analysis reporting blood group A associated with a substantially higher risk of COVID-19, [29] as well as a study from Turkey in which blood group A was the most frequently detected group among COVID-19 patients. [6]

Among comorbidities, hypertension was the most frequent, followed by diabetes, cardiac illness, and HIV/AIDS, a pattern supported by a study conducted in Ethiopia, [26] suggesting that patients with hypertension, followed by diabetes and cardiac illness, may be more likely to be infected.

Presence of complications and pregnancy showed significant associations with COVID-19 severity. Participants with complications were 14.96 times more likely to be in a more severe category than those without, holding other variables constant. [8] Pregnant participants were 85% less likely to be in a more severe category than non-pregnant participants, a finding that contrasts with a study conducted in Canada reporting pregnant women more likely to receive medical interventions related to severe COVID-19 infection, [28] a discrepancy that may warrant further investigation given the relatively small number of pregnant participants in this cohort.

### 4.1. Limitations and strengths

This study is strengthened by its use of an ordinal logistic regression model, appropriate for the ordered nature of the severity outcome. It is limited, however, by its single-center design, relatively small sample size (355), and predominantly urban catchment population. Data were drawn from patient charts rather than direct interview, which may not capture all relevant factors, and variables such as cultural considerations, occupation, and income, not examined in this study, may also influence COVID-19 severity.

In conclusion, this study found a substantial association between severity of COVID-19 infection and the occurrence of complications and pregnancy, but no association between ABO blood group and severity of COVID-19 pneumonia in this cohort. These findings support prioritizing rapid, aggressive management for pregnant patients and those developing complications in order to halt disease progression. They also suggest that ABO blood typing should not currently be regarded as a prognostic indicator in patients with COVID-19 in this setting, and that health authorities in Ethiopia may find greater value in directing resources toward comorbidity management, antenatal COVID-19 screening, and early recognition of complications than toward blood-group-based risk stratification.

## Data Availability

All data produced in the present study are available upon reasonable request to the authors

## ACKNOWLEDGMENTS

The authors thank the staff of Eka Kotebe General Hospital, COVID-19 Isolation and Treatment Center, for their support during data collection.

## DATA AVAILABILITY STATEMENT

The de-identified dataset analyzed for this study is available from the corresponding author on reasonable request, given the presence of patient-level clinical information collected under the ethical approval described above.

## AUTHORS’ ADDRESSES

Eden Bekele Urgecha, In-Patient Department, Eka Kotebe General Hospital, Addis Ababa, Ethiopia.

Fanuel Alemayehu Tefera, In-Patient Department, Eka Kotebe General Hospital, Addis Ababa, Ethiopia.

Zewdie Aderaw Alemu, General Public Health, Gamby Medical and Business College, Addis Ababa, Ethiopia.

Solomon Teshome Mekonen, School of Medicine, Mekelle University, Mekelle, Ethiopia, Addis Ababa, Ethiopia.

Etsub Bekele Orgecha, School of Pharmacy, Jimma University, Jimma, Ethiopia.

Saron Negasi Gidey, School of Medicine, Mekelle University, Mekelle, Ethiopia

Fiyameta Abraham Fissehatsion, School of Medicine, Mekelle University, Mekelle, Ethiopia.

